# Age-and Sex-Specific Distribution and Reference Values of Coronary Artery Calcium in a Large Asymptomatic Japanese Cohort

**DOI:** 10.1101/2025.09.11.25335612

**Authors:** Hidenobu Takagi, Masaharu Hirano, Takashi Asano, Kensuke Nishimiya, Taku Obara, Hideki Ota, Junichi Taguchi, Kei Takase

## Abstract

**Background:** The clinical use of coronary artery calcium (CAC) scoring for risk stratification in Japan is limited by the absence of population-specific reference data, as applying Western-derived thresholds is inappropriate due to known ethnic variations. This study aimed to establish the first comprehensive, age-and sex-specific CAC reference values for a healthy Japanese population.

**Methods:** In this single-center retrospective study, we analyzed data from 4,891 asymptomatic Japanese adults (63.2% men; median age, 58 years) without a history of ASCVD or diabetes. Age-and sex-specific CAC percentile curves were generated using nonparametric regression modeling.

**Results:** Men exhibited a higher CAC burden than women, with scores increasing with age in both sexes. The relationship between detectable CAC (CAC >0) and age was nonlinear: concave down for men and concave up for women, indicating different progression patterns. Compared to MESA data, the Japanese cohort had a markedly lower CAC burden than White participants and systematically lower scores than Chinese American women.

**Conclusions:** This study provides the first large-scale, age-and sex-specific CAC reference values for a healthy Japanese population. The generated percentile curves offer a practical tool for clinicians to immediately assess a patient’s CAC burden relative to their peers, underscoring that using foreign-derived thresholds is inappropriate for risk stratification in Japan.

## Introduction

Atherosclerotic cardiovascular disease (ASCVD) remains a leading cause of morbidity and mortality worldwide.^1^ Notably, approximately 50% of ASCVD-related deaths occur in asymptomatic individuals, highlighting the critical need for accurate risk stratification in asymptomatic populations and the timely initiation of preventive interventions in high-risk individuals.^2^ International guidelines recommend ASCVD risk assessment using prediction models, followed by lifestyle modification and preventive pharmacotherapy based on estimated risk.^3–5^ However, conventional risk models frequently overestimate or underestimate ASCVD risk,^6,7^ potentially leading to suboptimal therapeutic guidance. Although efforts have been made to refine existing models and develop novel predictive tools, risk stratification based solely on conventional parameters —such as age, sex, medical history, family history, and laboratory data— has inherent limitations. Additional modalities are needed to enhance risk prediction, particularly to appropriately reclassify individuals categorized as intermediate risk and to facilitate individualized preventive strategies.

Coronary artery calcification (CAC) is a well-established surrogate marker of subclinical atherosclerosis and provides incremental prognostic value beyond traditional risk factors.^8^ Numerous studies, including large-scale prospective cohorts, have demonstrated that CAC scoring significantly improves ASCVD risk prediction and enhances the reclassification of individuals with borderline or intermediate risk.^3,9,10^ In recognition of this evidence, the 2019 American College of Cardiology/American Heart Association (ACC/AHA) guidelines provide a Class IIa recommendation for the use of CAC scoring to supplement conventional risk assessment.^3^ In contrast, Japanese guidelines currently recommend risk assessment based solely on conventional factors and do not endorse CAC scoring for primary prevention, citing concerns about ethnic differences.^5^

The importance of population-specific reference values is well-documented. The Multi-Ethnic Study of Atherosclerosis (MESA) revealed substantial variations in CAC burden by ethnicity,^11^ and significant efforts have been made to establish reference values for various populations. Notably, a large-scale study in over 31,000 asymptomatic Koreans has provided robust data for that population,^12^ and a recent meta-analysis has summarized CAC reference ranges across multiple ethnicities, including some data from Japan.^13^ However, despite these crucial contributions, the direct application of such data to the Japanese population may be limited due to potential differences in lifestyle, diet, and genetic background, even within East Asian groups. Furthermore, existing data often lack the detailed, continuous percentile distributions necessary for a practical clinical tool that allows for the nuanced risk assessment of an individual patient relative to their peers. To date, a large-scale, contemporary reference standard tailored specifically for the Japanese population remains undefined.

Therefore, this study aimed to define the age-and sex-specific distribution of CAC in a large cohort of healthy Japanese individuals and to develop a practical reference tool with detailed percentile curves to support individualized ASCVD risk assessment and prevention in Japan.

## Materials and Methods

### Study Desing and Cohort

This single-center retrospective observational study was approved by the Ethics Committee of Tohoku University Graduate School of Medicine, Sendai, Japan (identifier: 2023-1-855). The requirement for written informed consent was waived due to the retrospective nature of the study. Asymptomatic individuals who underwent comprehensive health check-ups at Yamanakako Clinic (Yamanashi, Japan) between 2016 and 2023 were included. For individuals with multiple CAC examinations, only the earliest result was used. Exclusion criteria were the absence of CAC scanning or a history of percutaneous coronary intervention, myocardial infarction, stroke or transient ischemic attack, heart failure, or diabetes mellitus (**Figure 1**).

**Figure 1.**
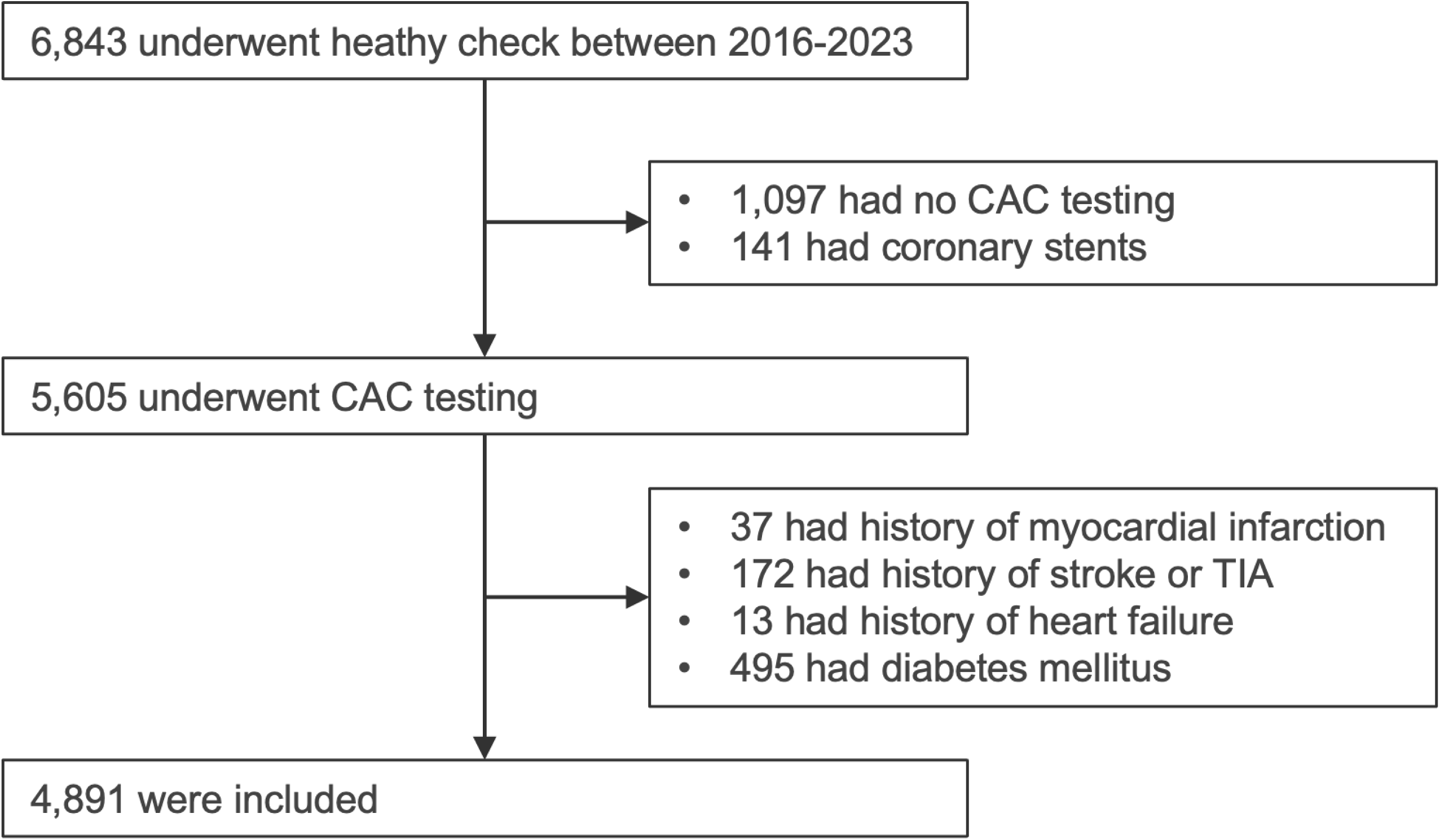
Flowchart of study participant selection. This flowchart illustrates the process of participant inclusion and exclusion, leading to the final cohort for analysis.

### Clinical Data Collection and Laboratory Measurements

Data on age, sex, medical history, antihypertensive and lipid-lowering medication use, and smoking status were obtained through a standardized questionnaire and verified using electronic medical records. Smoking status was categorized as “Never” (never smoked), “Past” (former smokers who had quit), or “Current” (actively smoking at the time of assessment).

Systolic and diastolic blood pressure were measured using an automated sphygmomanometer following at least three minutes of rest in a seated position. Blood samples were collected after an overnight fast, and serum lipid profiles (low-density lipoprotein cholesterol, high-density lipoprotein cholesterol, triglycerides) and hemoglobin A1c levels were analyzed using standardized enzymatic methods.

### Risk Stratification

The 10-year ASCVD risk was calculated for each participant using a validated risk prediction model derived from the Hisayama Study.^14^ Risk categories were defined as low risk (<5.0%), intermediate risk (5.0% to 19.9%), and high risk (≥20%), consistent with the American guideline.^3^

### Coronary Artery Calcium Scanning and Measurements

CAC scanning was performed using a standardized protocol with a 64-slice CT scanner (Biograph mCT Flow, Siemens Healthineers, Erlangen, Germany) without contrast and using electrocardiography-gated acquisition. CAC was quantified by experienced technicians using semiautomated software (AZE Virtual Place; AZE, Inc, Tokyo, Japan) according to the Agatston method ^15^. The analysis software was updated during the study period, with the Raijin module used until 2022 and the Class R module from 2023 onwards. CAC scores were recorded in the electronic medical record and categorized as follows: 0 (none), 1–10 (minimal), 11–100 (mild), 101–300 (moderate), 301–999 (severe), or >1000 (extensive).^16^

## Statistical Analysis

Continuous variables are presented as medians with interquartile ranges (25th–75th percentiles), and categorical variables as counts and percentages. The distribution of CAC scores in this cohort was highly right skewed for both men and women, as illustrated in **Supplementary Figure 1a**. To address this skewness, CAC was log-transformed as *log*(*CAC*+1) for all analyses involving continuous CAC values. The distribution of log-transformed CAC is shown in **Supplementary Figure 1b**. Following the methodology used in the Multi-Ethnic Study of Atherosclerosis (MESA) for establishing CAC percentiles,^17^ we first modelled the probability of detectable CAC (CAC >0) as a function of age, stratified by sex, using nonparametric locally weighted scatterplot smoothing (LOWESS) regression to account for the highly skewed distribution of CAC in asymptomatic populations. Next, we modelled the mean of *log*(*CAC*+1) as a function of age separately for each sex, using a smoothing span of 0.7. Residuals from this model were pooled, ranked, and used to calculate the 1st through 100th percentiles of the residual distribution. Finally, these percentiles were added to the fitted mean estimates to derive age-and sex-specific percentile values for CAC. All analyses were performed using R version 4.4.0.^18^

## Results

### Participant Characteristics

A total of 6,843 individuals underwent health check-ups between 2016 and 2023 and were screened for study eligibility (**Figure 1**). Of these, 1,097 individuals who did not undergo CAC testing and 141 individuals with coronary stents were excluded. Among the remaining 5,605 individuals, those with a history of myocardial infarction (n = 37), stroke or transient ischemic attack (n = 172), heart failure (n = 13), or diabetes mellitus (n = 495) were excluded. Consequently, 4,891 individuals were included in the final analysis. The characteristics of the study cohort are summarized in **Table 1**. The cohort was predominantly male (n = 3,094; 63.2%), with a median age of 58 years (interquartile range, 51–67). The most common age category among men was 45–54 years (n = 1,032; 33.3%), whereas among women, the 55–64 years age category was most prevalent (n = 669; 37.2%).

**Table 1.**
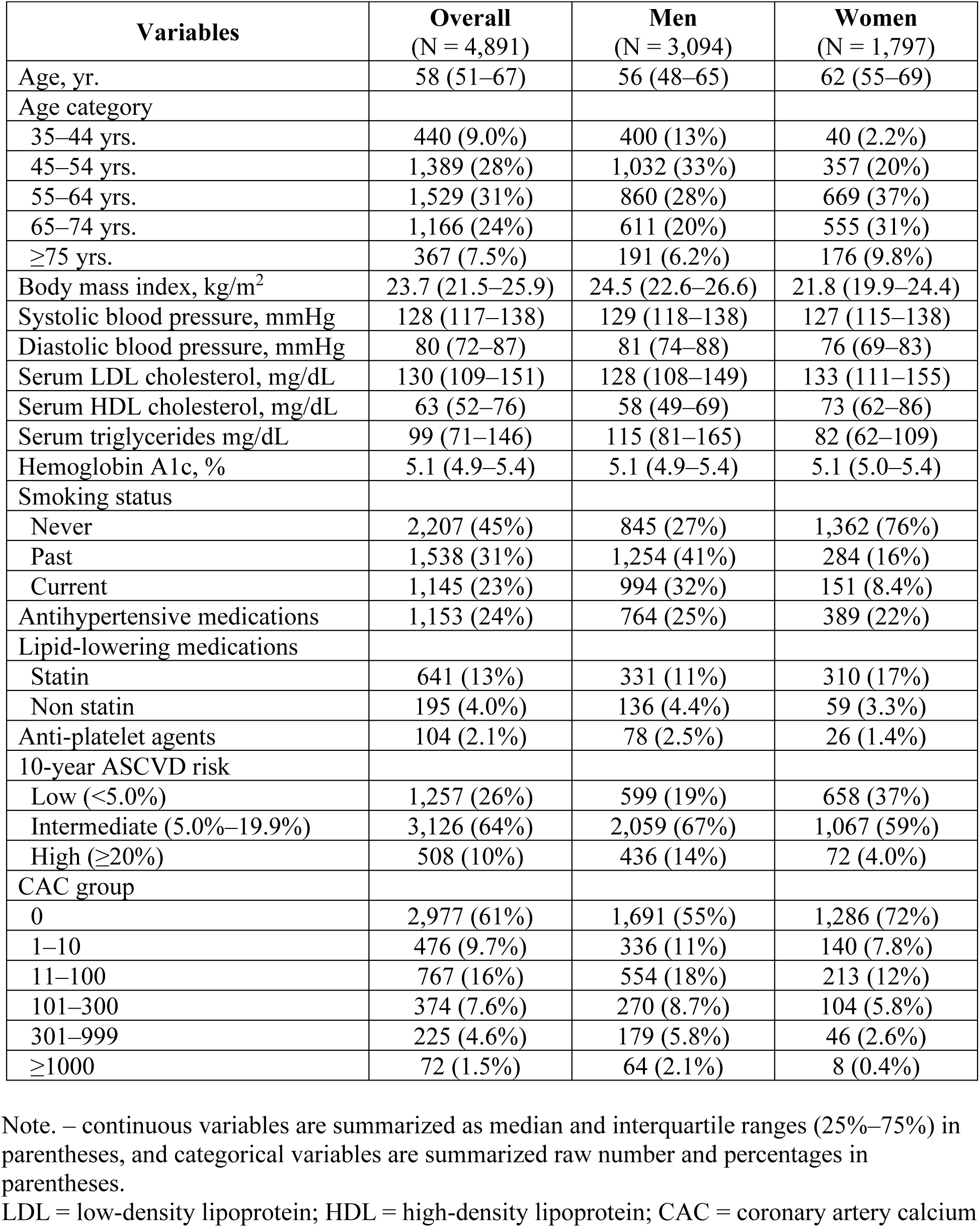
Characteristics of the Study Population.

| **Variables** | **Overall**<br>**(N = 4,891)** | **Men**<br>**(N = 3,094)** | **Women**<br>**(N = 1,797)** |
| --- | --- | --- | --- |
| Age, yr. | 58 (51–67) | 56 (48–65) | 62 (55–69) |
| Age category |  |  |  |
| 35–44 yrs. | 440 (9.0%) | 400 (13%) | 40 (2.2%) |
| 45–54 yrs. | 1,389 (28%) | 1,032 (33%) | 357 (20%) |
| 55–64 yrs. | 1,529 (31%) | 860 (28%) | 669 (37%) |
| 65–74 yrs. | 1,166 (24%) | 611 (20%) | 555 (31%) |
| ≥75 yrs. | 367 (7.5%) | 191 (6.2%) | 176 (9.8%) |
| Body mass index, kg/m<sup>2</sup> | 23.7 (21.5–25.9) | 24.5 (22.6–26.6) | 21.8 (19.9–24.4) |
| Systolic blood pressure, mmHg | 128 (117–138) | 129 (118–138) | 127 (115–138) |
| Diastolic blood pressure, mmHg | 80 (72–87) | 81 (74–88) | 76 (69–83) |
| Serum LDL cholesterol, mg/dL | 130 (109–151) | 128 (108–149) | 133 (111–155) |
| Serum HDL cholesterol, mg/dL | 63 (52–76) | 58 (49–69) | 73 (62–86) |
| Serum triglycerides mg/dL | 99 (71–146) | 115 (81–165) | 82 (62–109) |
| Hemoglobin A1c, % | 5.1 (4.9–5.4) | 5.1 (4.9–5.4) | 5.1 (5.0–5.4) |
| Smoking status |  |  |  |
| Never | 2,207 (45%) | 845 (27%) | 1,362 (76%) |
| Past | 1,538 (31%) | 1,254 (41%) | 284 (16%) |
| Current | 1,145 (23%) | 994 (32%) | 151 (8.4%) |
| Antihypertensive medications | 1,153 (24%) | 764 (25%) | 389 (22%) |
| Lipid-lowering medications |  |  |  |
| Statin | 641 (13%) | 331 (11%) | 310 (17%) |
| Non statin | 195 (4.0%) | 136 (4.4%) | 59 (3.3%) |
| Anti-platelet agents | 104 (2.1%) | 78 (2.5%) | 26 (1.4%) |
| 10-year ASCVD risk |  |  |  |
| Low (<5.0%) | 1,257 (26%) | 599 (19%) | 658 (37%) |
| Intermediate (5.0%–19.9%) | 3,126 (64%) | 2,059 (67%) | 1,067 (59%) |
| High (≥20%) | 508 (10%) | 436 (14%) | 72 (4.0%) |
| CAC group |  |  |  |
| 0 | 2,977 (61%) | 1,691 (55%) | 1,286 (72%) |
| 1–10 | 476 (9.7%) | 336 (11%) | 140 (7.8%) |
| 11–100 | 767 (16%) | 554 (18%) | 213 (12%) |
| 101–300 | 374 (7.6%) | 270 (8.7%) | 104 (5.8%) |
| 301–999 | 225 (4.6%) | 179 (5.8%) | 46 (2.6%) |
| ≥1000 | 72 (1.5%) | 64 (2.1%) | 8 (0.4%) |

Note. – continuous variables are summarized as median and interquartile ranges (25%–75%) in parentheses, and categorical variables are summarized raw number and percentages in parentheses.
LDL = low-density lipoprotein; HDL = high-density lipoprotein; CAC = coronary artery calcium

### Prevalence of Coronary Artery Calcium by Age and Sex

The estimated and observed prevalence of detectable coronary artery calcium (CAC score >0), mild to extensive CAC (CAC score ≥100), and severe to extensive CAC (CAC score ≥300) by age and sex are shown in **Figure 2**. In both men and women, the probability of detectable CAC increased with advancing age. The estimated probability curve for detectable CAC showed an upward convex shape in men, with a steep increase after approximately 40 years of age. In women, the curve was downward convex with a gradual increase observed after approximately 55 years of age. Among men, the estimated prevalence of CAC >0 exceeded 15% from approximately 45 years of age and surpassed 25% at approximately 50 years of age, with a progressive increase thereafter. In contrast, women exhibited a lower prevalence of detectable CAC across all age groups, with rates exceeding 15% around 55 years of age and surpassing 25% at approximately 60 years.

**Figure 2.**
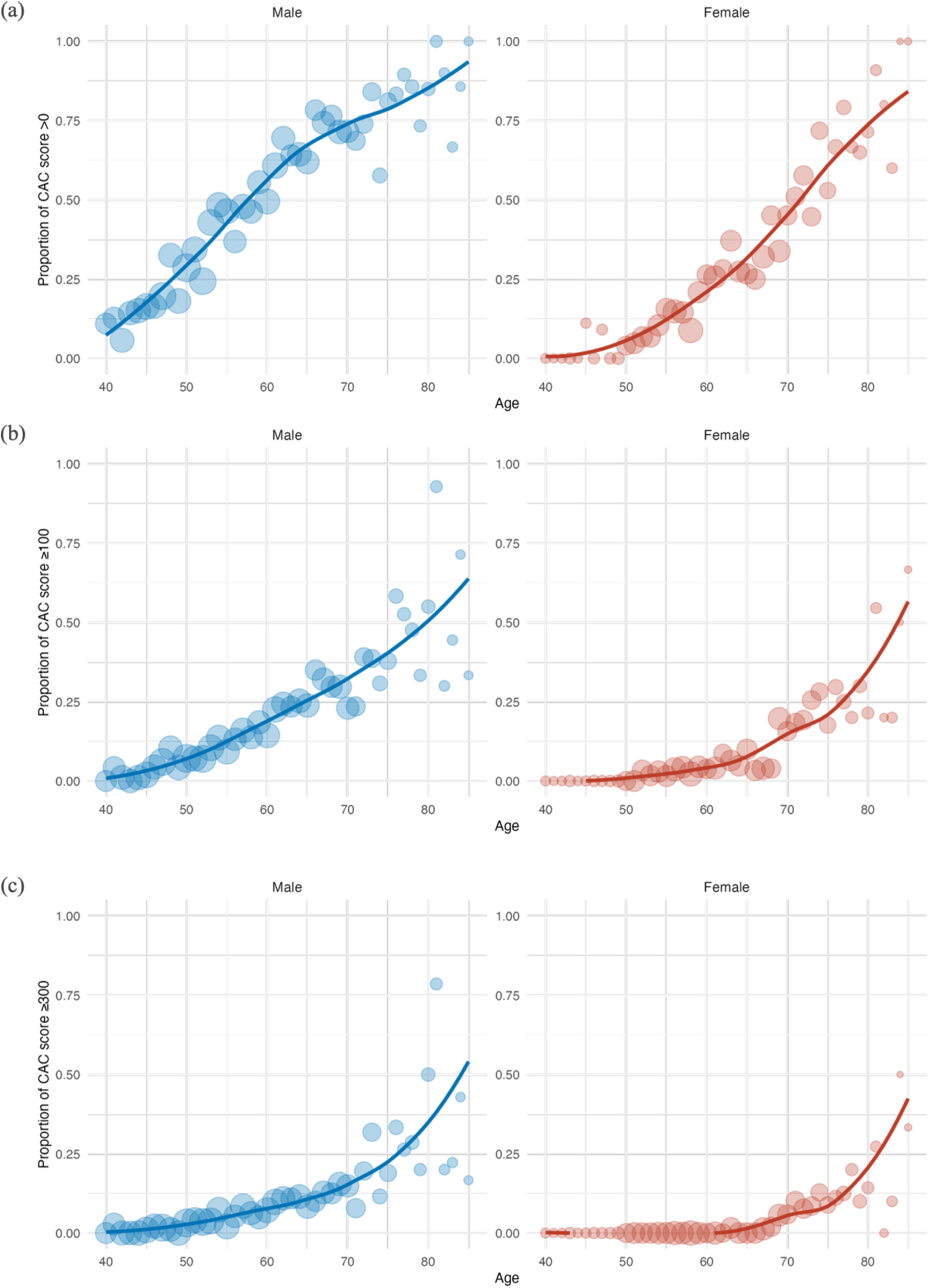
Estimated and observed probability of coronary artery calcium by age and sex. The graphs show the probability of (A) detectable CAC (score >0), (B) moderate or higher CAC (score ≥ 100), and (C) severe or higher CAC (score ≥300). Solid lines indicate the probability estimated by locally weighted scatterplot smoothing (LOWESS) regression. Shaded areas represent 95% confidence intervals. Dots represent the observed prevalence within 5-year age intervals.

Similarly, the prevalence of CAC ≥100 (**Figure 2b**) and CAC ≥300 (**Figure 2c**) increased with age in both sexes, with men consistently showing higher rates than women. For CAC ≥100, the estimated prevalence exceeded 15% in men from approximately 60 years of age, whereas in women, it remained below 15% until approximately 75 years of age. For CAC ≥300, the prevalence exceeded 15% in men after approximately 70 years, while in women, the prevalence surpassed 15% only after approximately 80 years of age.

### Distribution of Coronary Artery Calcium by Age and Sex

The estimated age-and sex-specific percentiles of CAC are shown in **Figure 3** and summarized numerically in **Table 2**. In both men and women, each estimated percentile of CAC increased exponentially with age. The increase was more pronounced in men, with higher CAC values observed across all age groups compared to women. Among men, the 75th percentile of CAC was greater than 0 in the 45–54 age group, reaching 55 in the 55–64 group, 140 in the 65– 74 group, and 441 in the ≥75 years group. In women, the 75th percentile remained 0 through the 55–64 age group, increasing to 20 in the 65–74 group and 137 in the ≥75 years group.

**Figure 3.**
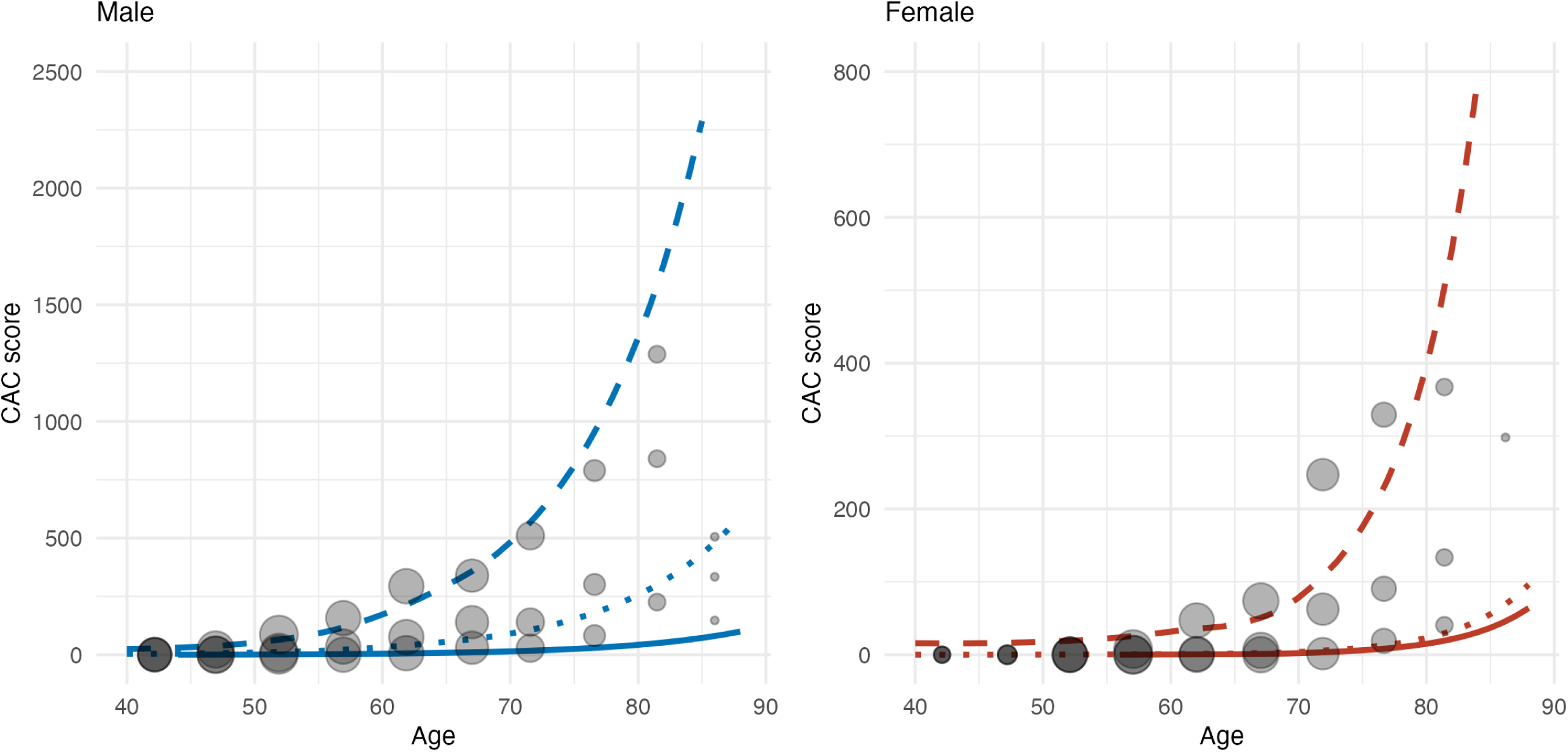
Estimated percentiles of coronary artery calcium score by age and sex. Each plot shows the estimated curves for the 50th, 75th, and 90th percentiles of the CAC score distribution across age for men (A) and women (B). The observed empirical percentiles for each 5-year age interval are overlaid as dots for reference.

**Table 2.** Estimated Percentile of CAC Score by Age Category, and Sex.

|  | **Age**<br>**35–44 yrs.** | **Age**<br>**45–54 yrs.** | **Age**<br>**55–64 yrs.** | **Age**<br>**65–74 yrs.** | **Age**<br>**≥75 yrs.** |
| --- | --- | --- | --- | --- | --- |
| **Male** |  |  |  |  |  |
| 25th percentile | 0 | 0 | 0 | 0 | 7 |
| 50th percentile | 0 | 0 | 2 | 29 | 106 |
| 75th percentile | 0 | 2 | 55 | 140 | 441 |
| 90th percentile | 1 | 58 | 237 | 407 | 1138 |
| 95th percentile | 14 | 151 | 430 | 823 | 1641 |
| **Female** |  |  |  |  |  |
| 25th percentile | 0 | 0 | 0 | 0 | 0 |
| 50th percentile | 0 | 0 | 0 | 0 | 31 |
| 75th percentile | 0 | 0 | 0 | 20 | 137 |
| 90th percentile | 0 | 0 | 23 | 131 | 410 |
| 95th percentile | 0 | 2 | 81 | 259 | 711 |

Note. – CAC = coronary artery calcium

## Discussions

In this large-scale study of asymptomatic Japanese individuals, we characterized the age-and sex-specific distribution of CAC scores. This study represents one of the largest analyses of CAC in a healthy Japanese population and provides reference percentiles to guide clinical interpretation. Our main findings are twofold. First, the prevalence of detectable CAC and CAC scores increased steadily with age in both sexes. Compared to women, men exhibited an earlier onset and higher burden of CAC across all age groups, indicating more accelerated subclinical atherosclerosis. Second, when compared to data from MESA, Japanese individuals demonstrated consistently lower CAC scores than their White counterparts across all age and sex groups.

Aging promotes vascular calcification through multiple pathways, including endothelial dysfunction, oxidative stress, chronic low-grade inflammation, arterial remodeling, and osteogenic transdifferentiation of vascular smooth muscle cells.^19–22^ In our cohort, men exceeded a 15% prevalence of detectable CAC (CAC >0) by age 45, whereas women reached this threshold approximately a decade later. A similar lag was observed for moderate (CAC ≥100) and severe (CAC ≥300) calcification, suggesting delayed vascular aging in women. These patterns are concordant with findings from MESA and other large cohorts,^17,23–26^ which consistently report an earlier onset and more rapid progression of calcification in men. The vasoprotective effects of estrogen are widely proposed to underlie these sex differences,^27^ with epidemiological data showing a sharp rise in coronary artery disease incidence in women following menopause. While the fundamental relationships between advancing age, male sex, and increased CAC appear universal across ethnicities, our findings underscore the need for Japanese-specific reference percentiles to account for systematically lower CAC scores relative to thresholds derived from Western cohorts.

Our study contributes to the growing body of literature defining CAC distributions in East Asian populations. A large-scale study in over 31,000 asymptomatic Koreans has provided robust reference data,^12^ and a recent meta-analysis summarized CAC ranges across multiple ethnicities, including some from Japan. Our results are broadly consistent with these studies, confirming a lower CAC burden in East Asians compared to Western populations. However, we also observed significant heterogeneity within Asian subgroups. As illustrated in **Figure 4**, which situates our cohort within the context of MESA’s multi-ethnic data, Japanese women in our cohort exhibited systematically lower CAC scores than Chinese American women in MESA, a finding that highlights potential differences in lifestyle, diet, or genetic factors that are not captured by broad ethnic categories. This observation is further supported by studies like MASALA, which found that South Asians have a CAC burden comparable to or higher than White Americans.^29^ These findings reinforce that “Asian” is not a monolithic category for ASCVD risk. Our results also align with previous, smaller-scale Japanese studies, such as the Shiga Epidemiological Study, which noted a lower CAC burden in Japanese men that diverged from US White men with age.^30^ The unique contribution of our study is the provision of detailed, continuous percentile distributions from a large, contemporary Japanese cohort. This offers a more granular and clinically applicable tool for individual risk assessment than was previously available. By allowing clinicians to place a patient’s CAC score in the context of their direct peers, our reference curves strengthen the call for population-specific thresholds to avoid the potential misclassification of risk that may arise from applying foreign-derived or generalized Asian data.

**Figure 4.**
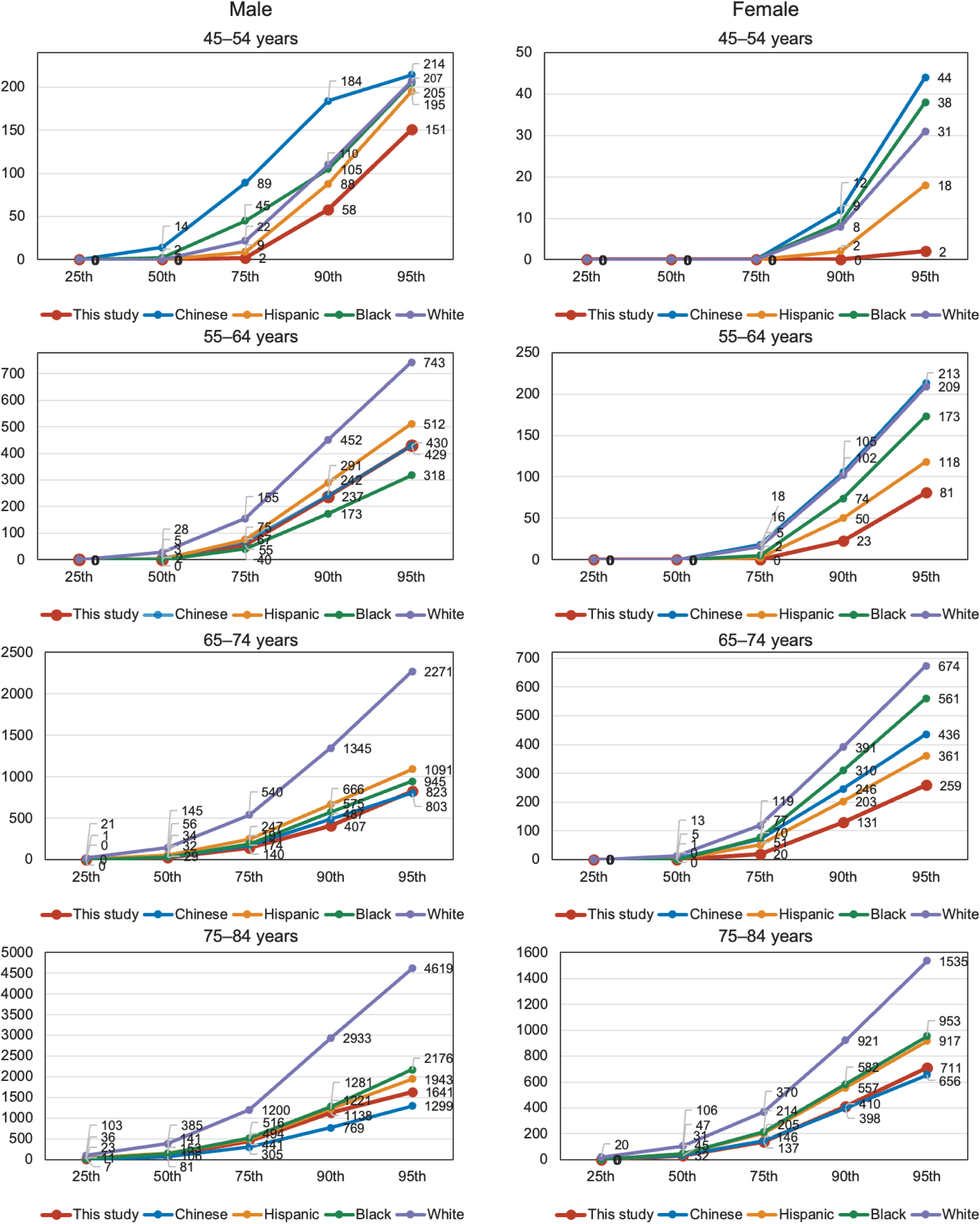
Comparison of median coronary artery calcium scores among different ethnic groups. The CAC scores for Japanese men and women from this study are compared with those of White, Black, Hispanic, and Chinese American participants from the Multi-Ethnic Study of Atherosclerosis (MESA)^17^. This figure highlights the differences in CAC burden across ethnicities.

This study has several limitations. First, the single-center design, using data from a health check-up facility, may limit the generalizability of our findings to the entire Japanese population. Participants in such programs may be more health-conscious or have a different socioeconomic status compared to the general public, potentially introducing selection bias. Second, the study’s cross-sectional nature precludes the assessment of CAC progression over time or the prognostic value of these CAC distributions for predicting future ASCVD events. We have established reference values, but their ability to predict outcomes in a Japanese context needs validation in prospective cohort studies. Third, although data were collected systematically, the retrospective design relies on the accuracy of existing electronic medical records and self-reported questionnaires, which may be subject to information bias. Finally, while our cohort was ethnically Japanese, the findings may not apply to other ethnic minority groups residing in Japan. Despite these limitations, this study provides the largest and most robust dataset to date for CAC reference values in a healthy Japanese population.

In conclusion, this study established comprehensive age-and sex-specific reference values for coronary artery calcium in a large cohort of asymptomatic Japanese individuals. Our data reveal a substantially lower CAC burden in the Japanese population compared to Western cohorts and highlight key differences even within East Asian subgroups, particularly for women. These findings underscore the inadequacy of applying foreign-derived CAC thresholds to Japanese patients and provide an essential, population-specific tool to improve the accuracy of ASCVD risk stratification in Japan. Future prospective studies are warranted to validate the prognostic significance of these Japanese-specific CAC percentiles for predicting long-term cardiovascular outcomes.

## Abbreviations

ASCVD: atherosclerotic cardiovascular disease
CAC: coronary artery calcium
CT: computed tomography
MESA: Multi-Ethnic Study of Atherosclerosis

## Data Availability Statement

The data that support the findings of this study are not publicly available due to ethical restrictions protecting patient privacy.

## Notes

**Funding:** This work was supported by research funding from the Department of Early Imaging Diagnosis (a joint research course); by a Grant-in-Aid for Young Scientists from the Japan Society for the Promotion of Science (JSPS KAKENHI Grant Number 23K15150); and by the Young Investigator Collaborative Research Encouragement Grant from Tohoku University Graduate School of Medicine (Fiscal Year 2023).

